# Availability of evidence and comparative effectiveness for surgical versus drug interventions: an overview of systematic reviews

**DOI:** 10.1101/2023.01.30.23285207

**Authors:** Emmanuel A. Zavalis, Anaïs Rameau, Anirudh Saraswathula, Joachim Vist, Ewoud Schuit, John P. A. Ioannidis

## Abstract

**Objectives:** To examine the prevalence of comparisons of surgery to drug regimens, the strength of evidence of such comparisons, and whether surgery or the drug intervention was favored.

**Design:** Systematic review of systematic reviews (umbrella review)

**Data sources:** Cochrane Database of Systematic Reviews (CDSR)

**Eligibility criteria and synthesis of results:** Using the search term “surg*” in CDSR, we retrieved systematic reviews of surgical interventions. Abstracts were subsequently screened to find systematic reviews that aimed to compare surgical to drug interventions; and then, among them, those that included any randomized controlled trials (RCTs) for such comparisons. Trial results data were extracted manually and synthesized into random-effects meta-analyses.

**Results:** Overall, 188 systematic reviews intended to compare surgery versus drugs. Only 41 included data from at least one RCT (total, 165 RCTs with data) and covered a total of 103 different outcomes of various comparisons of surgery versus drugs. A GRADE assessment was performed by the Cochrane reviewers for 87 (83%) outcomes in the reviews, indicating the strength of evidence was high in 4 outcomes (4%), moderate in 22 (21%), low in 27 (26%) and very low in 33 (32%). Based on 95% confidence intervals, the surgical intervention was favored in 38/103 (37%), and the drugs were favored in 13/103 (13%) outcomes. Of the outcomes with high GRADE rating, only one showed conclusive superiority (sphincterotomy was better than medical therapy for anal fissure). Of the 22 outcomes with moderate GRADE rating, 6 (27%) were inconclusive, 14 (64%) were in favor of surgery, and 2 (9%) were in favor of drugs.

**Conclusions:** Though the relative merits of surgical versus drug interventions are important to know for many diseases, high strength randomized evidence is rare. More randomized trials comparing surgery to drug interventions are needed.

**Protocol registration:** https://osf.io/p9x3j

**Funding:** The work of John Ioannidis has been funded by an unrestricted gift from Sue and Bob O’Donnell. Anaïs Rameau is supported by a Paul B. Beeson Emerging Leaders Career Development Award in Aging (K76 AG079040) from the National Institute on Aging and by the Bridge2AI award (OT2 OD032720) from the NIH Common Fund. Anirudh Saraswathula is supported by the National Institute on Deafness and Other Communication Disorders training grant 2T32DC000027.

**Financial disclosure:** Anaïs Rameau is a medical advisor for Perceptron Health, Inc.

**Summary boxes:** 

**Section 1: What is already known on this topic:**

- Many conditions and diseases can be managed either with surgery or with drugs. Comparative effectiveness of different treatment options is important to know for shared decision-making.

**Section 2: What this study adds:**

- An assessment of the entire Cochrane Database of Systematic Reviews found 188 reviews that intended to assess surgical versus drug interventions, but only 41 had at least one randomized trial.
- Only four of the 103 assessed outcomes had high strength of evidence according to GRADE assessments.
- More evidence is needed to compare the relative merits of surgical and drug interventions and sequestration of these major modes of interventions should be overcome in clinical trial agendas.

## Introduction

Many diseases are treated or managed with surgery. Some may also be addressed by pharmaceutical interventions, and studying the effectiveness of these different interventions is important in optimizing shared decision-making for patients and physicians. However, the amount and certainty of the evidence we hold in healthcare is limited[1], and this situation might be worse for surgical interventions due to serious challenges in running placebo-controlled or comparative effectiveness trials[2]. Challenges to controlled trials include unique patient anatomy, operator dependent variables such as the skill or experience of the surgeon[3–5], and the difficulty of successful blinding[6]. Due to these challenges, randomized controlled trials (RCTs) in surgery are less common than in non-surgical medical specialties. Although there have been calls to strengthen the quality of the evidence in surgery[2,7,8], these challenges have resulted in relatively few RCTs assessing surgical interventions, particularly in comparison to medical treatments.

A summary of the existing body of evidence on surgical versus medical interventions across diseases does not exist in the literature. To synthesize this existing body of evidence is of paramount importance to evidence-based care and informed decisions in the clinic where surgery or drugs are available interventions. To find RCTs comparing surgical vs. pharmaceutical interventions, we set up to conduct an umbrella review (an overview of systematic reviews) [9,10] by searching the Cochrane Database of Systematic Reviews for reviews considering comparisons of surgery to drugs, analyze the strength of the evidence and evaluate results of these comparisons. Finally, we explored whether results favoring surgery were more likely to be published in the surgical literature.

## Methods

### Search strategy and selection criteria

We queried the Cochrane Database of Systematic Reviews using the term “surg*” in “Title/Abstract/Keywords” on April 25, 2022. Inclusion criteria for reviews were consideration of RCTs and comparing a surgical to a drug intervention.

A surgical intervention was defined as a procedural technique aiming to change anatomy to treat or alleviate a pathology or symptom (including dermatological excisions). We excluded endoscopic and endovascular procedures since many of them are performed by medical rather than surgical specialists. A drug intervention was defined as a treatment that utilized a non-supplement and non-vitamin, pharmaceutical agent. Dental procedures, radiation treatment, as well as comparisons of surgery vs. no treatment or only placebo were excluded from our study. Cochrane reviews that intended to compare surgical and pharmaceutical interventions were considered even in cases where the review was unsuccessful in finding any such comparisons.

As many surgical procedures also require drug regimens (e.g., pre-operatively or as background treatment), we allowed comparisons where the surgical arm including a drug intervention was compared to a drug intervention as well. Comparisons of surgery to surgery plus drugs were not eligible, as both arms used surgery.

The articles’ abstracts were reviewed by EAZ, and JV who coded the reviews independently for eligibility and then sought to reach a consensus. Remaining differences were mediated by JPAI.

### Main outcomes

The main outcomes assessed in this umbrella review were the number of Cochrane systematic reviews that considered comparisons of surgical and drug interventions, and the number of systematic reviews that found any eligible RCTs comparing a surgical and a drug arm. The strength of evidence of the existing comparison was also treated as a main outcome, as were the direction of effects in the review assessments, both in the original Cochrane analysis and our standardized re-analysis).

### Data extraction

EAZ extracted data for the included systematic reviews. In the cases where no data were available from any RCT, the review was tabulated into the category of reviews that found no evidence from RCTs on comparisons of surgical versus drug treatments.

The systematic reviews were classified into their corresponding surgical specialty field: cardiac surgery, dermatology, general surgery, neurosurgery, obstetrics and gynaecology, ophthalmology, orthopaedic surgery, otolaryngology, plastic surgery, thoracic surgery, urology and vascular surgery.

Whenever data were available from at least one RCT comparing a surgical to a drug arm, we identified the primary outcome(s) of the systematic review for the eligible comparison(s) by examining the methods section of the systematic review. Data, in the form of contingency tables or means, standard deviations and number of participants in each arms, from individual RCTs were then collected from Cochrane eligible reviews. The primary outcomes were classified as mortality, composite, and non-mortality. We also collected information on GRADE assessments for the eligible comparisons and outcomes and the summary effect size as well as the 95% confidence interval of the effect for the eligible comparison outcomes.

### Meta-analysis

Cochrane reviewers may have used different statistical models in each topic to combine the results of RCTs in meta-analyses. To achieve standardization, we recalculated the summary effect size and heterogeneity for each topic using a random effects model following the Hartung-Knapp-Sidik-Jonkman approach[11,12] so that all outcomes/topics would be analyzed with the same statistical methods. The modified Haldane-Anscombe continuity correction was used, i.e. when studies had no event in either the surgical or the drug arm we added 0.5 to the entire contingency table of the specific study[13].

The analysis of the data was performed using R version 4.1.3 (2022-03-10), with assessment of statistical significance being using a threshold for α of 0.005, as previously proposed[14]. The Wilson approach was used for confidence intervals (99.5%) created for the primary outcomes.

### Additions to the protocol

The original pre-registered protocol can be found in *https://osf.io/p9x3j*. Some additions were made during the process of conducting this umbrella review. For each systematic review, we noted the search date to understand how old they might be. We also extracted the year of publication of each RCT to capture how recent the evidence was. Finally, we extracted the specialty orientation of the journal, in which the RCT was published, using the categories “mostly surgical”, “general”, and “mostly non-surgical”. The category “mostly surgical” includes those journals that have “surgery” in their title, those that have the name of a surgical specialty in their title, and those affiliated with a surgical society. The category “general” pertains to journals that cover all of medicine and its specialties, surgical and non-surgical. The category “mostly non-surgical” includes all the remaining journals. We assessed whether the direction of effects (favoring surgery or favoring drug) was associated with the type of journal, hypothesizing that RCTs published in mostly surgical journals may be more likely than other journals to favor surgery.

## Results

### Search results

The selection flowchart for Cochrane systematic reviews is represented in Figure 1. The search strategy retrieved 2495 articles from the Cochrane Database of Systematic Reviews. Among them, 440 were excluded by an automated search for withdrawn reviews and of studies with no mention of the word surgery and any of its variations in the abstract. Further manual inspection of titles and abstracts resulted in 223 Cochrane reviews being potentially eligible. Upon full-text evaluation, 35 were excluded: in 5 reviews, the surgical and drug treatments were not in separate arms and hence they were not an eligible head-to-head comparison[15–19]; in 7 reviews, there was no surgical intervention arm[20–26]; in 17 reviews, there was no drug intervention [27– 32,32–42]); 2 reviews were excluded for evaluating an endoscopic intervention [43,44]; 3 reviews were excluded for evaluating an endovascular intervention [45–47]; and finally 1 review was excluded for being an umbrella review[48].

**Figure 1.**
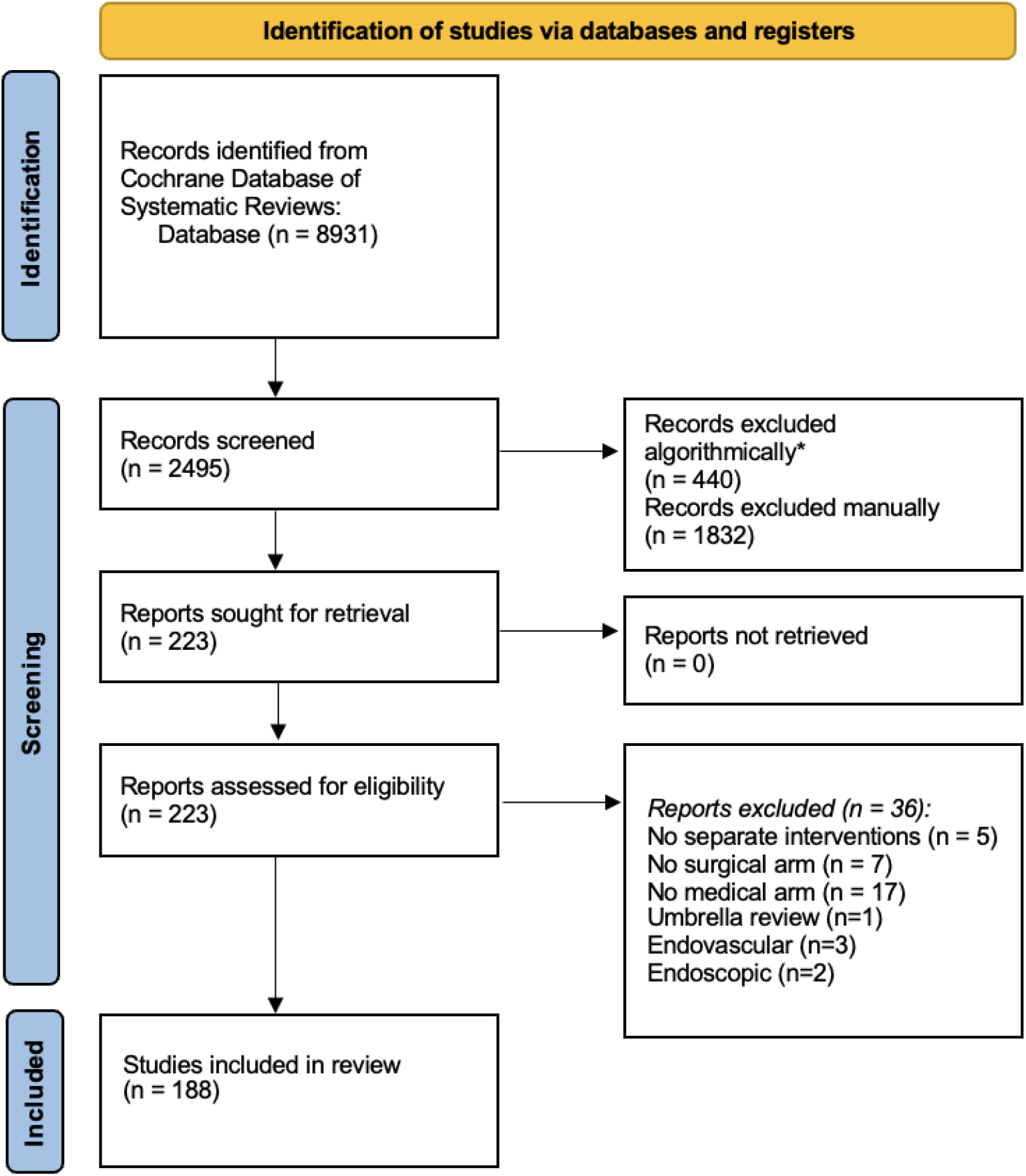
PRISMA study selection flowchart *Searched title and abstract for withdrawals, and for abstracts without the word surgery or any of its variations to exclude them

Therefore, 188 Cochrane reviews were found to meet the inclusion criteria (Supplement 1). Of those, 147 Cochrane reviews aimed to investigate surgical versus drug interventions but were unable to find any RCTs meeting their selection criteria. The remaining 41 reviews contained data for at least one RCT in at least one head-to-head comparison of a surgical versus a drug intervention arm (22% (99.5% CI 14 to 31%)).

The 188 reviews covered all major surgical specialties (Supplementary Table 1), with the most commonly represented specialties being general surgery (n=35), obstetrics and gynecology (n=31), ophthalmology (n=25), orthopedic surgery (n=23) and otolaryngology (n=23). No significant difference was found across specialties in the proportion of reviews that contained data from at least one RCT for a surgery versus drug comparison (Fisher’s exact p=0.62).

### Comparative treatment effect for surgery versus drug comparisons

The 41 eligible reviews with data included 103 comparisons of surgery versus drug treatments with data on various primary outcomes (Table 1), and they included data from a total of 165 RCTs with a total of 295 primary outcome assessments. For the 165 trials, the median publication year was 2005 and the interquartile range was 1994 to 2016. The median search date year of the eligible reviews was 2016 (interquartile range, 2010 to 2022).

**Table 1.**
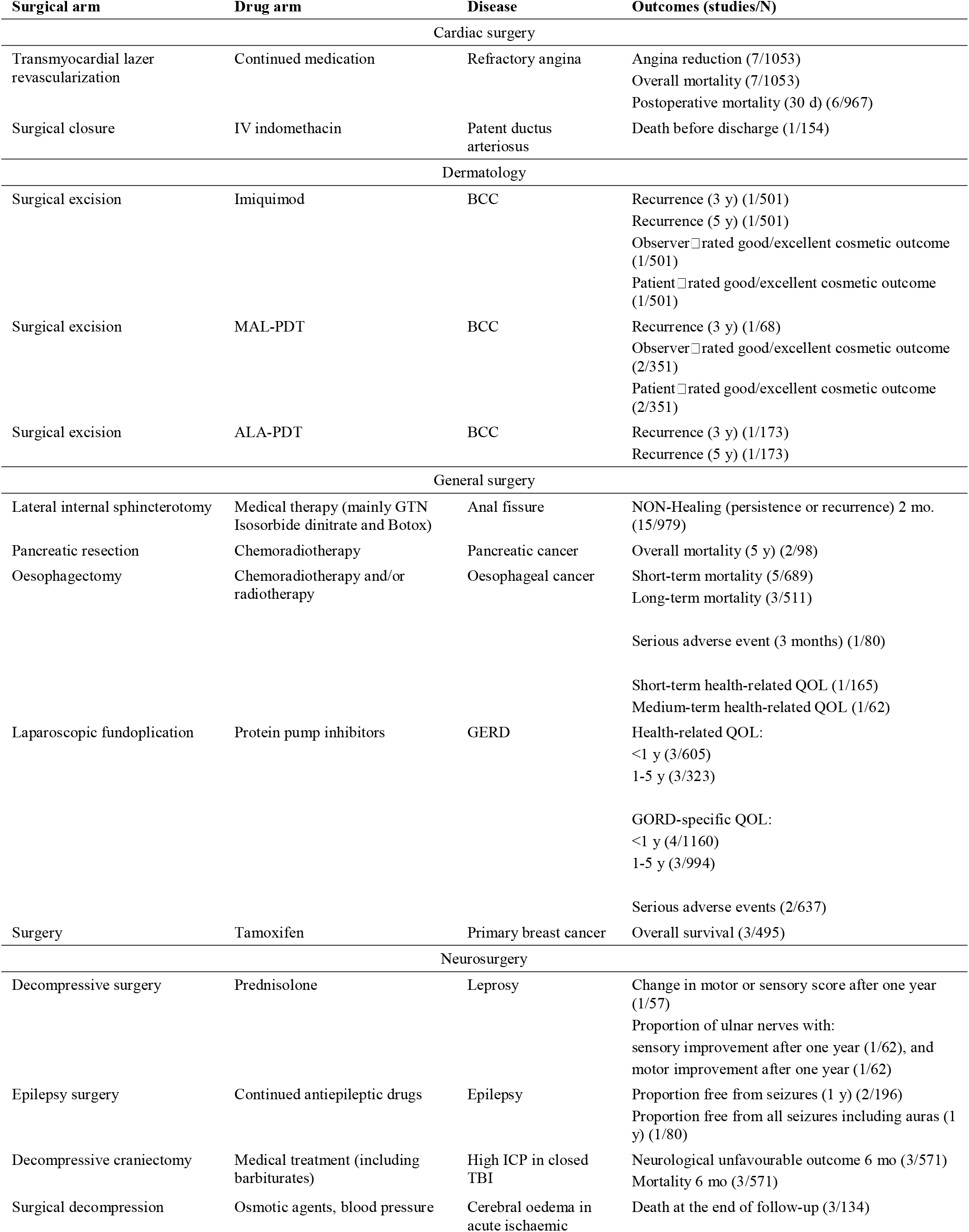
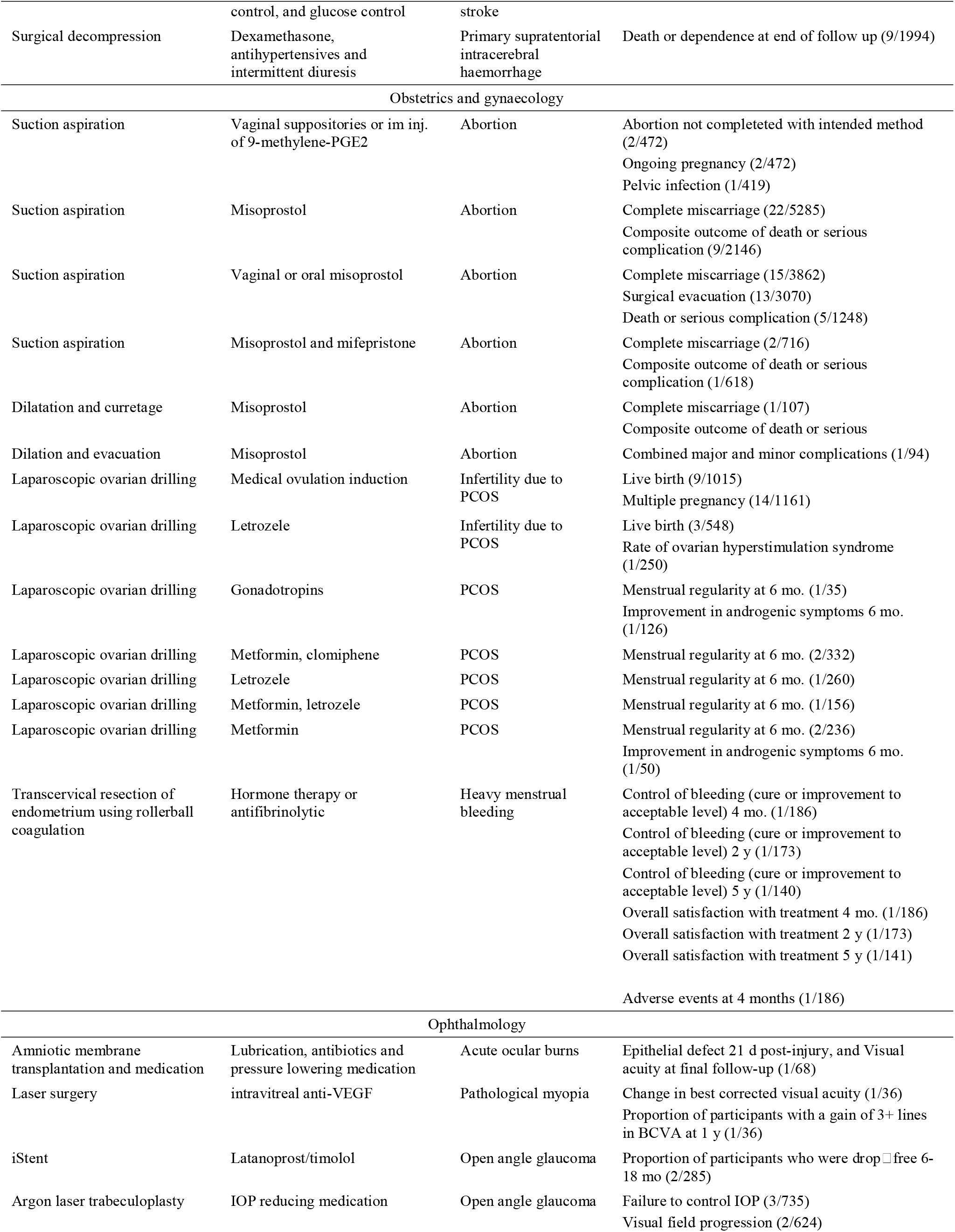
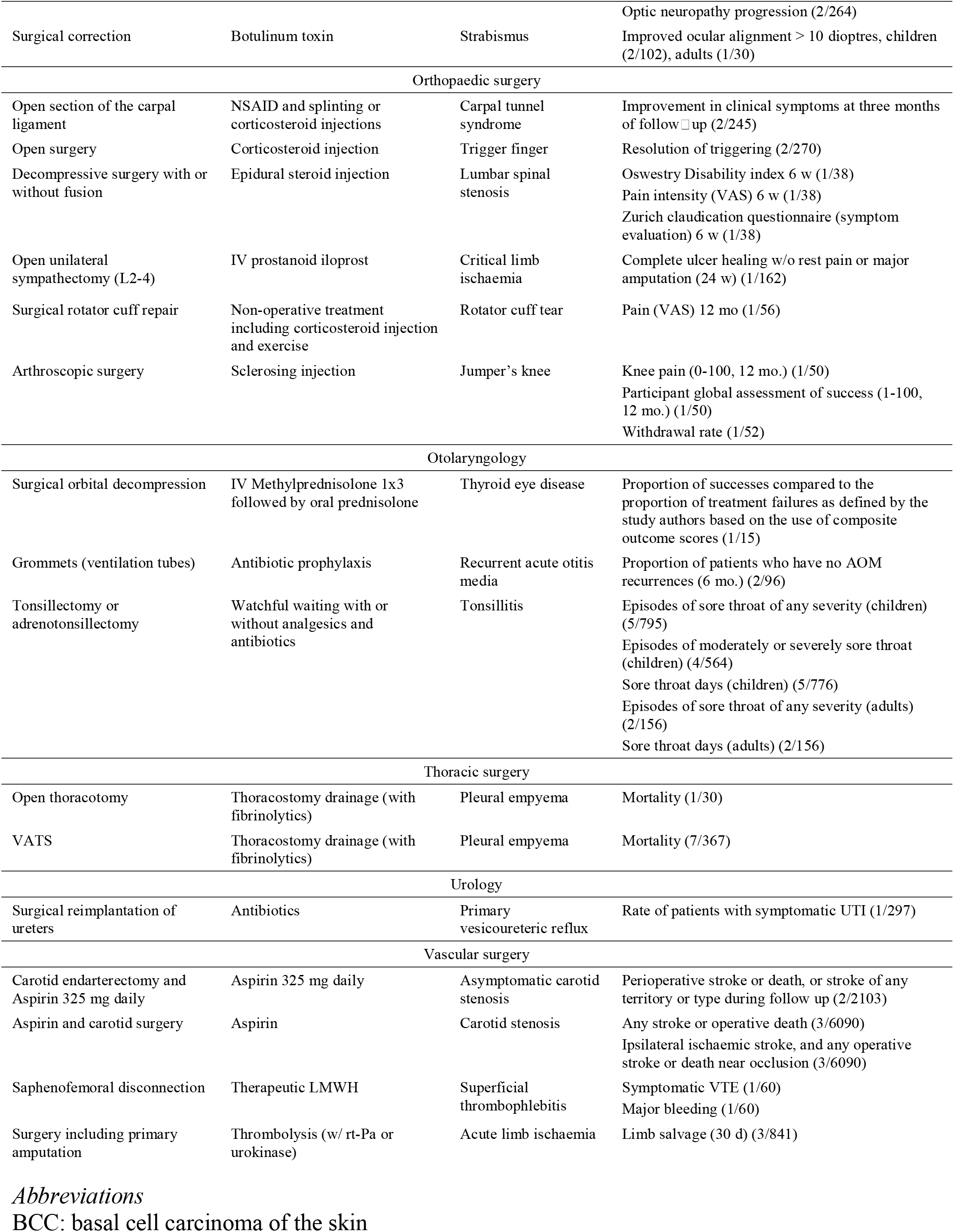
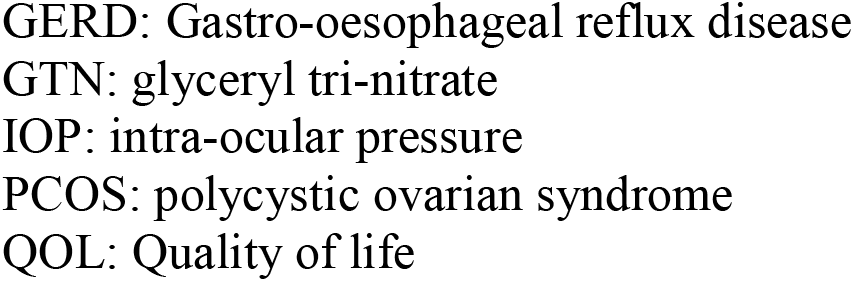
Eligible comparisons of surgical versus medical interventions

Based on the 95% confidence interval of the summary estimate obtained by the Cochrane review authors, surgery was more effective in 36 of the 103 outcomes of various comparisons (35% (99.5% CI 23 to 49%)), and drugs were more effective in 15 (15% (99.5% CI 6 to 26%)). Fifty two (50% (99.5 CI% 37 to 64%)) outcomes were inconclusive. The respective numbers were 1/12 (8%), 1/12 (8%), and 10/12 (83%) for mortality outcomes; 3/11 (27%), 3/11 (27%) and 5/11 (46%) for composite outcomes; and 32/80 (40%), 11/80 (14%), and 37/80 (46%) for non-mortality outcomes.

When we standardized the meta-analyses to use the same random effects method for all analyses, surgery was favored in 28/103 outcomes (32%), drugs were favored in 9 (10%) outcomes and 66 (58%) outcomes were inconclusive. The respective numbers were 1/12 (8%), 0/12 (0%), and 11/12 (92%) for mortality outcomes; 3/11 (18%), 2/11 (27%) and 6/11 (55%) for composite outcomes; and 24/80 (30%) 7/80 (9%), and 49/80 (61%) for non-mortality outcomes.

Table 2 shows the topics for which the surgical intervention was found to be more effective and Table 3 shows those where the drug arm was found to be more effective, all according to the Cochrane authors’ analysis. Supplementary Table 2 does the same for the topics for which the comparisons were inconclusive.

**Table 2.**
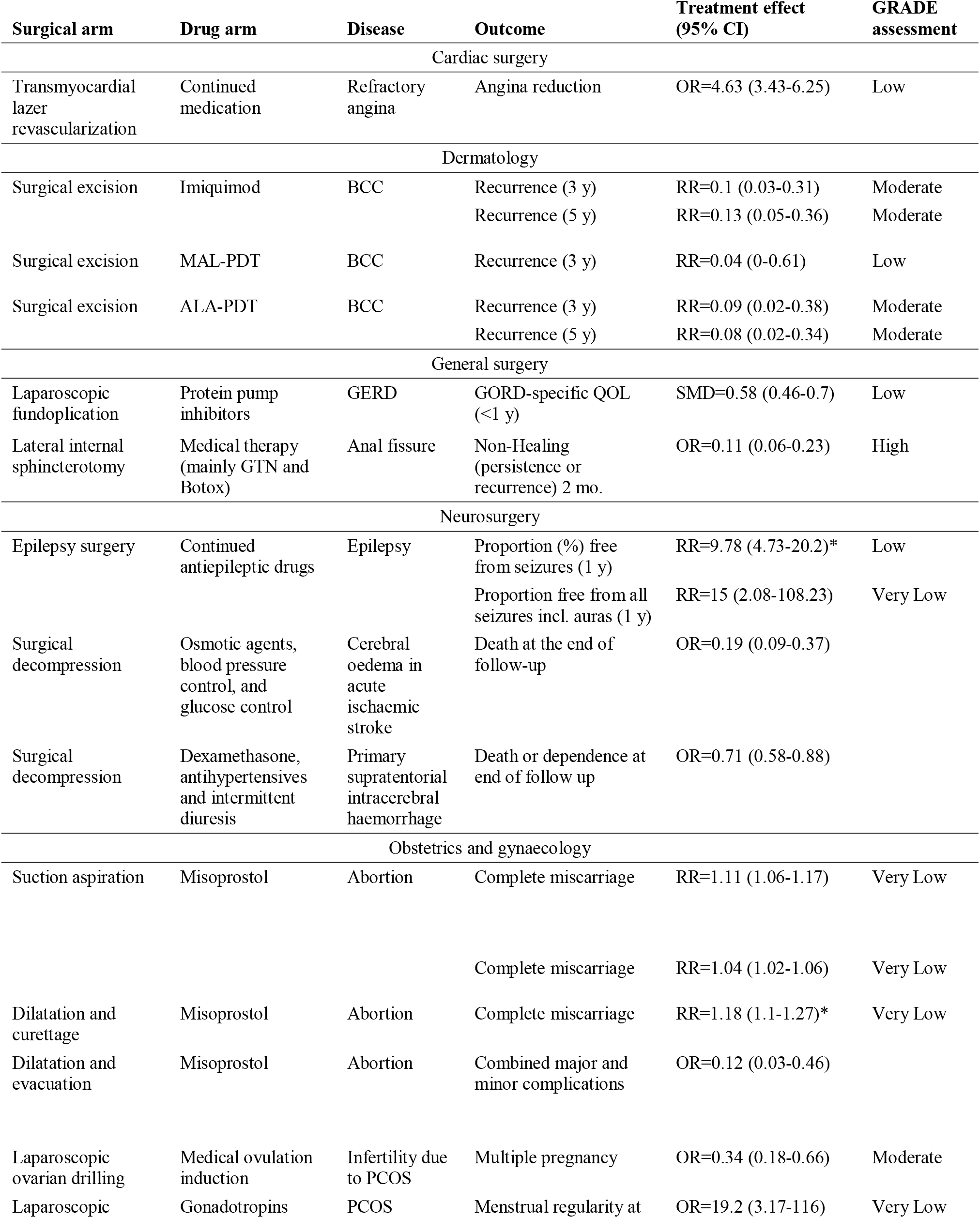
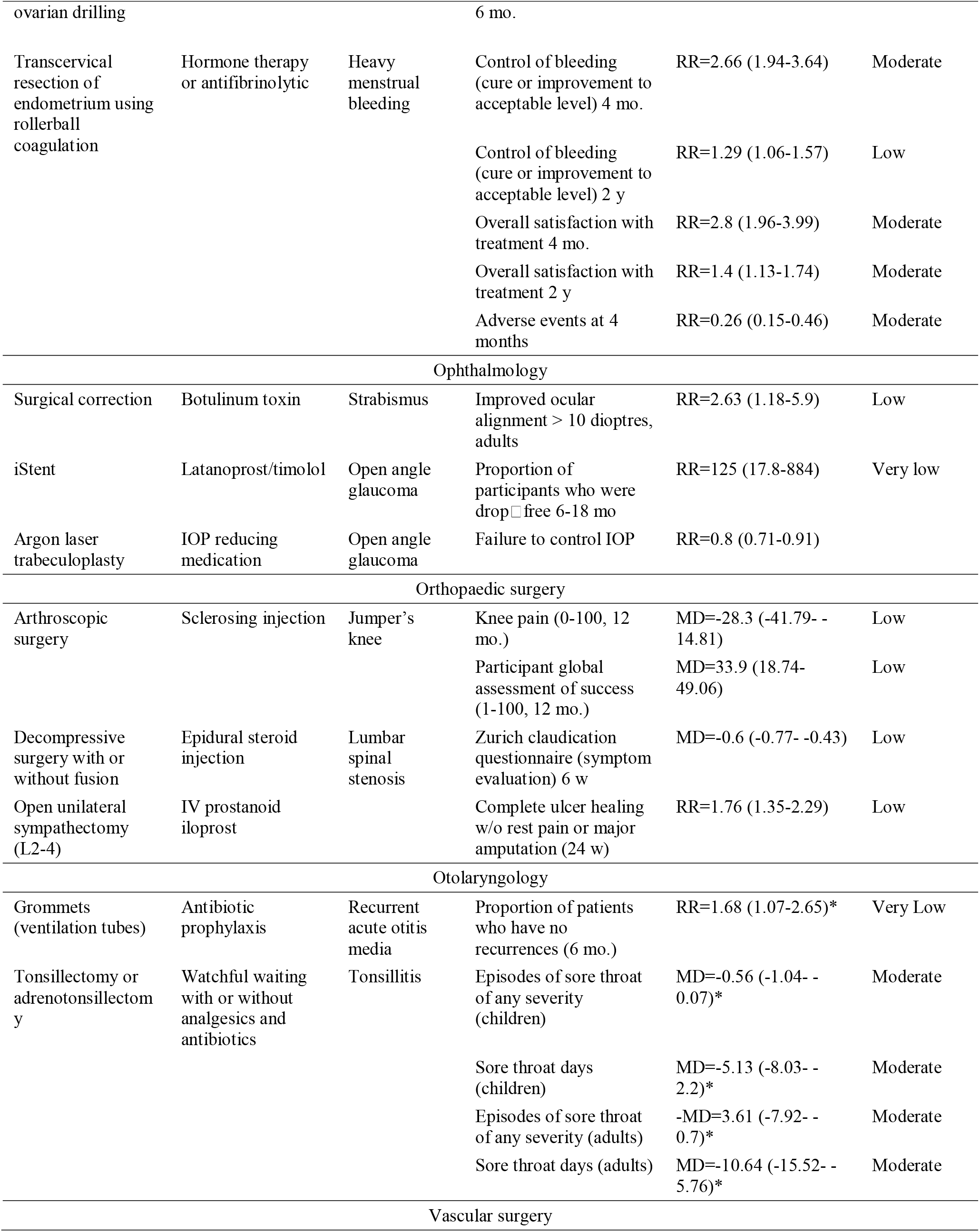
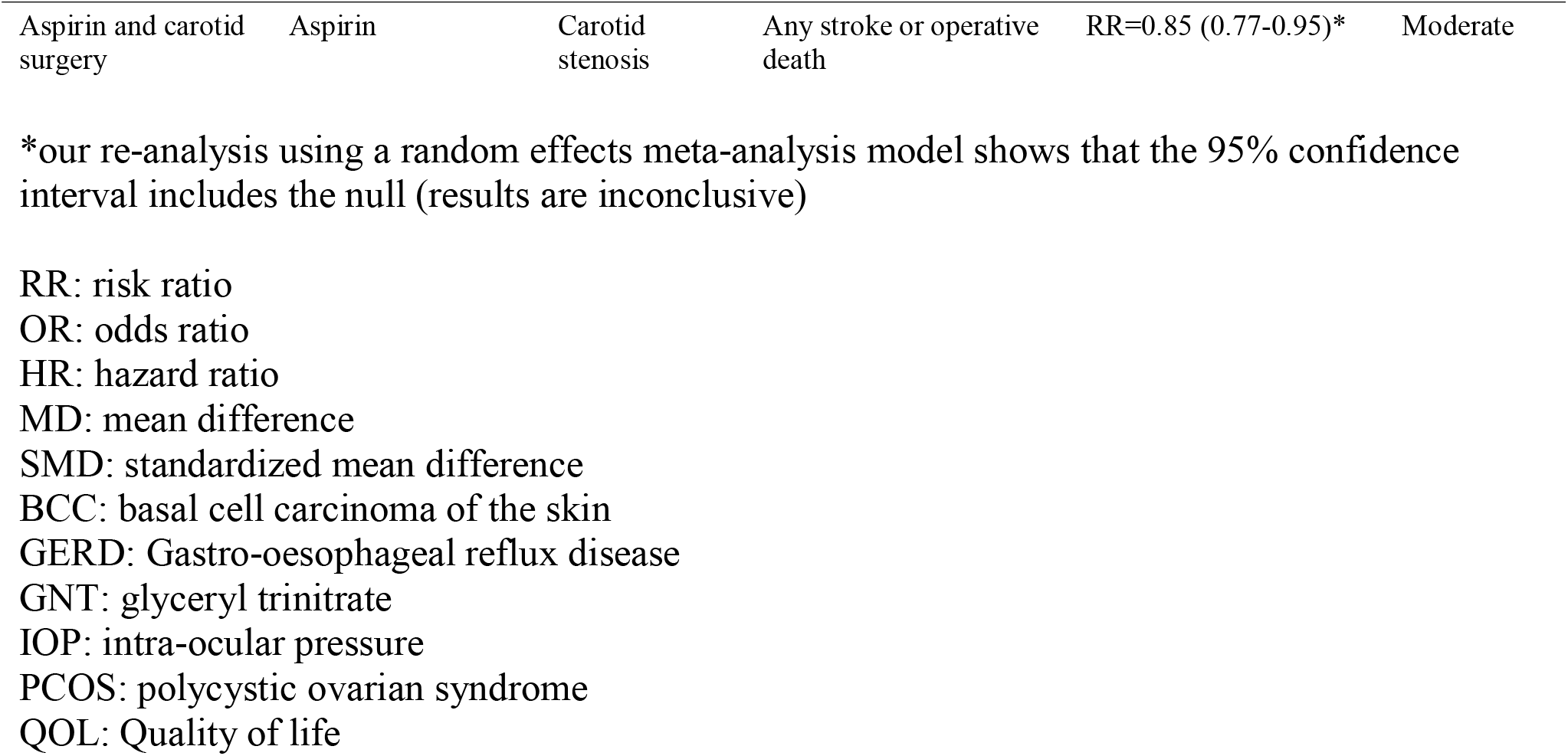
Comparisons where the surgical treatment was superior to the drug treatment

**Table 3.**
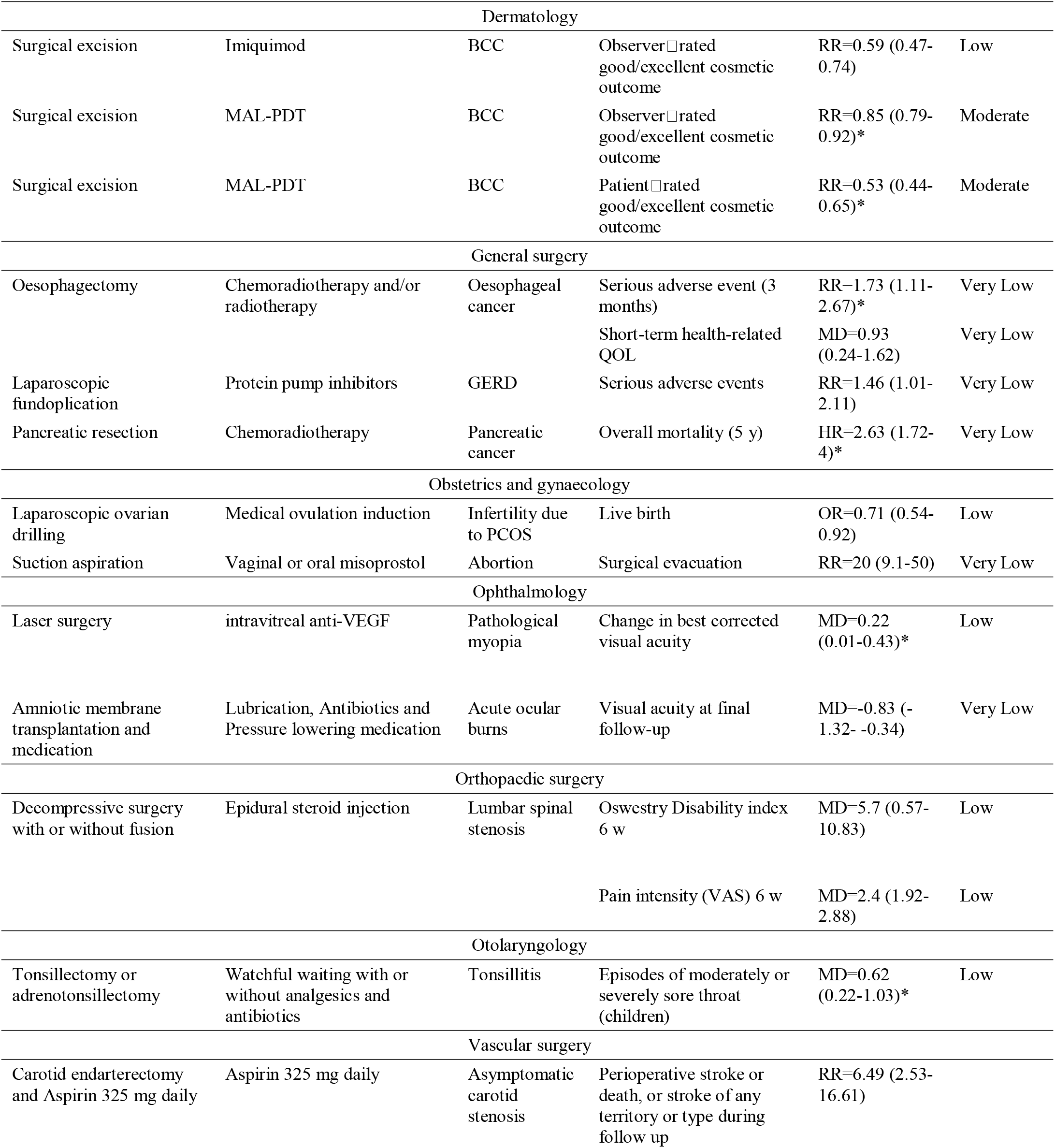
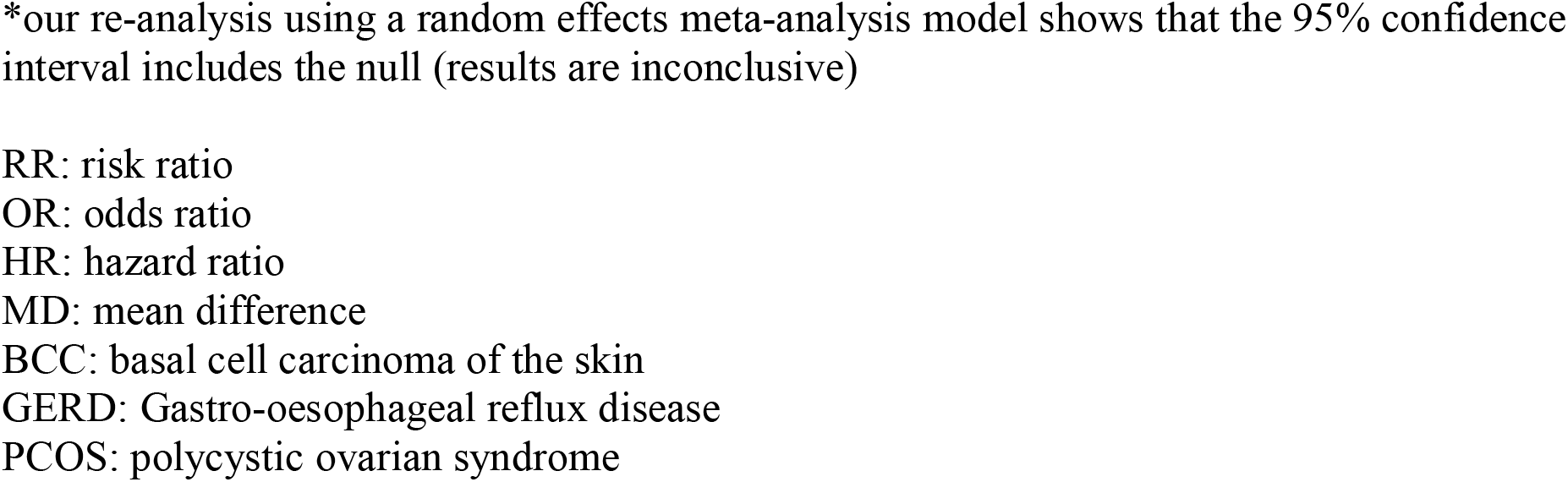
Comparisons where the drug treatment was superior to the surgical treatment

| Surgical arm | Drug arm | Disease | Outcome | Treatment effect (95% CI) | GRADE assessment |
| --- | --- | --- | --- | --- | --- |
| Dermatology |  |  |  |  |  |
| Surgical excision | Imiquimod | BCC | Observer-rated good/excellent cosmetic outcome | RR=0.59 (0.47-0.74) | Low |
| Surgical excision | MAL-PDT | BCC | Observer-rated good/excellent cosmetic outcome | RR=0.85 (0.79-0.92)* | Moderate |
| Surgical excision | MAL-PDT | BCC | Patient-rated good/excellent cosmetic outcome | RR=0.53 (0.44-0.65)* | Moderate |
| General surgery |  |  |  |  |  |
| Oesophagectomy | Chemoradiotherapy and/or radiotherapy | Oesophageal cancer | Serious adverse event (3 months) | RR=1.73 (1.11-2.67)* | Very Low |
|  |  |  | Short-term health-related QOL | MD=0.93 (0.24-1.62) | Very Low |
| Laparoscopic fundoplication | Protein pump inhibitors | GERD | Serious adverse events | RR=1.46 (1.01-2.11) | Very Low |
| Pancreatic resection | Chemoradiotherapy | Pancreatic cancer | Overall mortality (5 y) | HR=2.63 (1.72-4)* | Very Low |
| Obstetrics and gynaecology |  |  |  |  |  |
| Laparoscopic ovarian drilling | Medical ovulation induction | Infertility due to PCOS | Live birth | OR=0.71 (0.54-0.92) | Low |
| Suction aspiration | Vaginal or oral misoprostol | Abortion | Surgical evacuation | RR=20 (9.1-50) | Very Low |
| Ophthalmology |  |  |  |  |  |
| Laser surgery | intravitreal anti-VEGF | Pathological myopia | Change in best corrected visual acuity | MD=0.22 (0.01-0.43)* | Low |
| Amniotic membrane transplantation and medication | Lubrication, Antibiotics and Pressure lowering medication | Acute ocular burns | Visual acuity at final follow-up | MD=-0.83 (-1.32- -0.34) | Very Low |
| Orthopaedic surgery |  |  |  |  |  |
| Decompressive surgery with or without fusion | Epidural steroid injection | Lumbar spinal stenosis | Oswestry Disability index 6 w | MD=5.7 (0.57-10.83) | Low |
|  |  |  | Pain intensity (VAS) 6 w | MD=2.4 (1.92-2.88) | Low |
| Otolaryngology |  |  |  |  |  |
| Tonsillectomy or adenotonsillectomy | Watchful waiting with or without analgesics and antibiotics | Tonsillitis | Episodes of moderately or severely sore throat (children) | MD=0.62 (0.22-1.03)* | Low |
| Vascular surgery |  |  |  |  |  |
| Carotid endarterectomy and Aspirin 325 mg daily | Aspirin 325 mg daily | Asymptomatic carotid stenosis | Perioperative stroke or death, or stroke of any territory or type during follow up | RR=6.49 (2.53-16.61) |  |

\*our re-analysis using a random effects meta-analysis model shows that the 95% confidence interval includes the null (results are inconclusive)
RR: risk ratio
OR: odds ratio
HR: hazard ratio
MD: mean difference
BCC: basal cell carcinoma of the skin
GERD: Gastro-oesophageal reflux disease
PCOS: polycystic ovarian syndrome

### Strength of evidence according to GRADE

GRADE assessment of the strength of the evidence showed high rating for 4 outcomes (4%), moderate for 22 (21%), low for 27 (26%), and very low for 33 (32%). No GRADE assessment was performed for 17 (17%) outcomes.

According to GRADE assessments, only cardiac surgery, obstetrics and gynecology and general surgery interventions had high GRADE ratings. Otolaryngology and dermatology had many moderate ratings. Almost all other GRADE ratings were low or very low (Table 4).

**Table 4.** GRADE assessment across specialties

| **Specialty** | **Very Low** | **Low** | **Moderate** | **High** | **None available** |
| --- | --- | --- | --- | --- | --- |
| Cardiac surgery | 0 (0) | 1 (25) | 0 (0) | 2 (50) | 1 (25) |
| Dermatology | 0 (0) | 3 (33) | 6 (67) | 0 (0) | 0 (0) |
| General surgery | 9 (69) | 3 (23) | 0 (0) | 1 (8) | 0 (0) |
| Neurosurgery | 5 (50) | 2 (20) | 1 (10) | 0 (0) | 2 (20) |
| Obstetrics and gynecology | 14 (45) | 4 (13) | 7 (23) | 1 (3) | 5 (16) |
| Ophthalmology | 2 (20) | 5 (50) | 0 (0) | 0 (0) | 3 (30) |
| Orthopaedic surgery | 2 (20) | 6 (60) | 1 (10) | 0 (0) | 1 (10) |
| Otolaryngology | 1 (14) | 1 (14) | 4 (57) | 0 (0) | 1 (14) |
| Thoracic surgery | 0 (0) | 1 (50) | 1 (50) | 0 (0) | 0 (0) |
| Urology | 0 (0) | 0 (0) | 0 (0) | 0 (0) | 1 (100) |
| Vascular surgery | 0 (0) | 1 (17) | 2 (33) | 0 (0) | 3 (50) |

Of the four outcomes with high GRADE rating, sphincterotomy for anal fissure showed superiority over medical treatment while the other three comparisons were inconclusive. Of the 22 outcomes with moderate GRADE rating, 6 (27%) were inconclusive, 14 (64%) were in favor of surgery, and 2 (9%) were in favor of the drug regimen according to the calculations of the Cochrane authors (14 (64%), were inconclusive, 7 (32%) favored the surgical arm and 1 (5%) were in favor of the drug regimen according to our standard random-effects calculations).

### Results of RCTs according to journal of publication

Of the 165 eligible RCTs (295 outcome assessments), 73 RCTs (133 assessments) were published in mostly surgical journals, 38 RCTs (69 assessments) in general journals, and 54 RCTs (93 assessments) in mostly non-surgical journals. Based on 95% confidence intervals for the assessments of RCTs published in mostly surgical journals, 40/133 (30%) were in favor of surgery, 14/133 (11%) were in favor of drugs, and 79/133 (59%) were inconclusive. The respective numbers for the assessments of RCTs published in general journals were 27/69 (39%), 5/69 (7%), and 37/69 (53%); and for the assessments of RCTs published in mostly non-surgical journals they were 22/93 (24%), 15/93 (16%), and 56 (60%), respectively. The proportion of RCTs favoring surgery was not significantly higher in mostly surgical journals (30%) compared to other journals (39% and 24% for general and non-surgical journals respectively) (p=0.18 by Fisher’s exact test).

## Discussion

### Main findings

In a subset of Cochrane reviews that aimed to compare surgery to drugs we found that only 1 in 5 systematic reviews that had shown interest in such comparisons eventually found data from any RCTs for comparisons of the two modes of interventions. Furthermore, the majority of the comparisons where RCTs of surgery versus drugs had inconclusive results, and also had low or very low strength of the evidence on GRADE assessments. Anal fissure was the only disease in our sample that had high GRADE evidence and a direction of effect indicating that one intervention (sphincterotomy) was more effective. Consequently, in the vast majority of cases where surgical and pharmaceutical interventions are available for treatment, an evidence-based decision in the clinic is difficult. Our secondary post hoc analysis of the type of journal where the eligible RCTs were published showed that results published in surgical journals were not necessarily more prone to favor the surgical arm of an RCT over the pharmaceutical arm.

### Strengths

This study covers the entire Cochrane database which is considered a high-quality comprehensive collection of systematic reviews. Cochrane reviews tend to address questions typically asked in routine clinical practice and underpin many clinical guideline recommendations, making this sample all the more relevant to everyday practice [49]. Another strength of this study is that all surgical specialties were included. This is, therefore, to our knowledge the first project aiming to assess the extent of comparative evidence for surgery versus pharmacotherapy for a diverse spectrum of diseases.

### Limitations

Our analysis has several limitations. First, our pre-defined inclusion criteria excluded non-pharmacological medical interventions. Several comparisons may be found in the literature where surgery is compared against non-surgical non-pharmacological medical interventions, such as CPAP or radiotherapy. We also excluded endovascular and endoscopic procedures since they may be performed by surgical and medical specialists. These eligibility choices aimed to achieve some homogeneity in a project that is by definition already very heterogeneous. The use of an algorithm to filter out papers with no mention of the word surgery as well as the search strategy itself may have led to us missing reviews that discuss a particular surgical procedure but never explicitly mention the word surgery but merely the name of the intervention.

Second, we focused exclusively on RCTs, but other types of evidence, e.g., non-randomized controlled trials, or uncontrolled clinical trials may also exist and sometimes their results may be compelling enough to deem a randomized study unnecessary. Such unquestionable superiority in the absence of randomized evidence is however unlikely [50]. Efforts such as IDEAL [8] have laid out much of the groundwork for performing RCTs in surgical research, yet a dearth of RCTs in the surgical realm of research persists to this day.

Third, only one database (Cochrane Database of Systematic Reviews) was used for this study and we did not examine non-Cochrane meta-analyses published as journal articles. While the database aims to be all-inclusive, there are still some topics in medical and surgical care that have not been covered by Cochrane reviews.

Fourth, it is possible that within the same disease, subgroups of patients may be eligible only for medical or only for surgical treatment, or that one or the other approach is much better only for specific subgroups. With the dearth of evidence we found for the overall analysis, identification of such subgroup effects would be unlikely and error-prone.

### Context of these findings

Sequestration between different disciplines and specialties[51] may lead to isolation of specialists which use different tools, and this may lead to a lack of comparisons of the treatments that each specialty uses. Each specialty may have its own community, journals, meetings, and research agenda, limiting communication between different specialists even though they may be dealing with the same disease from different angles and with different therapeutic sets. This lack of communication may also be due to differences in mentorship and the trend of sub-specialization in medical training separating clinicians and their practices even further [52], or to differing incentive structures.

Prior literature comparing surgical and medical interventions has assessed specific treatments, such as that for basal cell carcinoma[51], and demonstrated that sequestration was prominent. Despite a large number of trials, almost all of them compared medical interventions among themselves, or surgicalaggressive interventions among themselves, rather than comparing between these two groups of treatment even though both groups of treatment could have been used. Our work shows that this issue of sequestration is widespread in surgical vs. pharmaceutical interventions.

### Implications

This study suggests that comparisons of pharmaceutical and surgical interventions are infrequent. Even accepting the difficulties in performing RCTs involving surgical interventions, our results still indicate a need for more comparative effectiveness research and for improved communication between surgical and medical specialties to bridge this gap in evidence. There are, of course, barriers to this. Head-to-head comparisons of treatments are often disfavored by manufacturers leery of jeopardizing their product against that of a competitor [53,54], and incentives unfortunately exist for both surgical and medical practitioners to promote treatments they are able to offer. Moving forward, both medical and surgical professional societies should collaborate to design fair and unbiased trials, and funders should also keep such research on their radars to try and overcome these structural obstacles.

### Future research

Future clinical research should try to expand the scope, volume, and methodological rigor of comparative evidence on surgical versus medical interventions. This work should involve both surgical and medical specialists and should also incorporate patient preferences. Long-term patient-centered outcomes, including both benefits and harms should become available to put surgical and medical practices into proper perspective.

## Supporting information

Supplement 1-List of included studies

## Data Availability

The PRISMA checklists, code, and data as well as the protocol are found in the Open Science Framework repository and is found in https://osf.io/p9x3j

## Authors’ contribution

The draft of the manuscript and the analysis was initially outlined and drafted by EAZ and JPAI with input from all authors in iterations. All authors had access to data, and approved the project design and critically reviewed, contributed to the final version of this work, and approve it.

## Transparency statement

Authors declare manuscript is an honest, accurate, and transparent account of the study being reported; no important aspects of the study have been omitted; and that any discrepancies from the study as originally planned (and, if relevant, registered) have been explained in the published protocol.

## Ethics approval

No ethics approval was required

## Supplementary material

**Supplementary table 1.** Reviews per specialty

| **Specialty** | **Total reviews** | **Reviews with at least one comparison (%)** |
| --- | --- | --- |
| Cardiac surgery | 6 | 2 (33) |
| Dermatology | 5 | 1 (20) |
| General surgery | 35 | 5 (14) |
| Neurosurgery | 12 | 5 (42) |
| Obstetrics and gynecology | 31 | 8 (26) |
| Ophthalmology | 25 | 5 (20) |
| Orthopaedic surgery | 23 | 6 (26) |
| Otolaryngology | 23 | 3 (13) |
| Thoracic surgery | 9 | 1 (11) |
| Urology | 7 | 1 (14) |
| Vascular surgery | 12 | 4 (33) |

**Supplementary Table 2.**
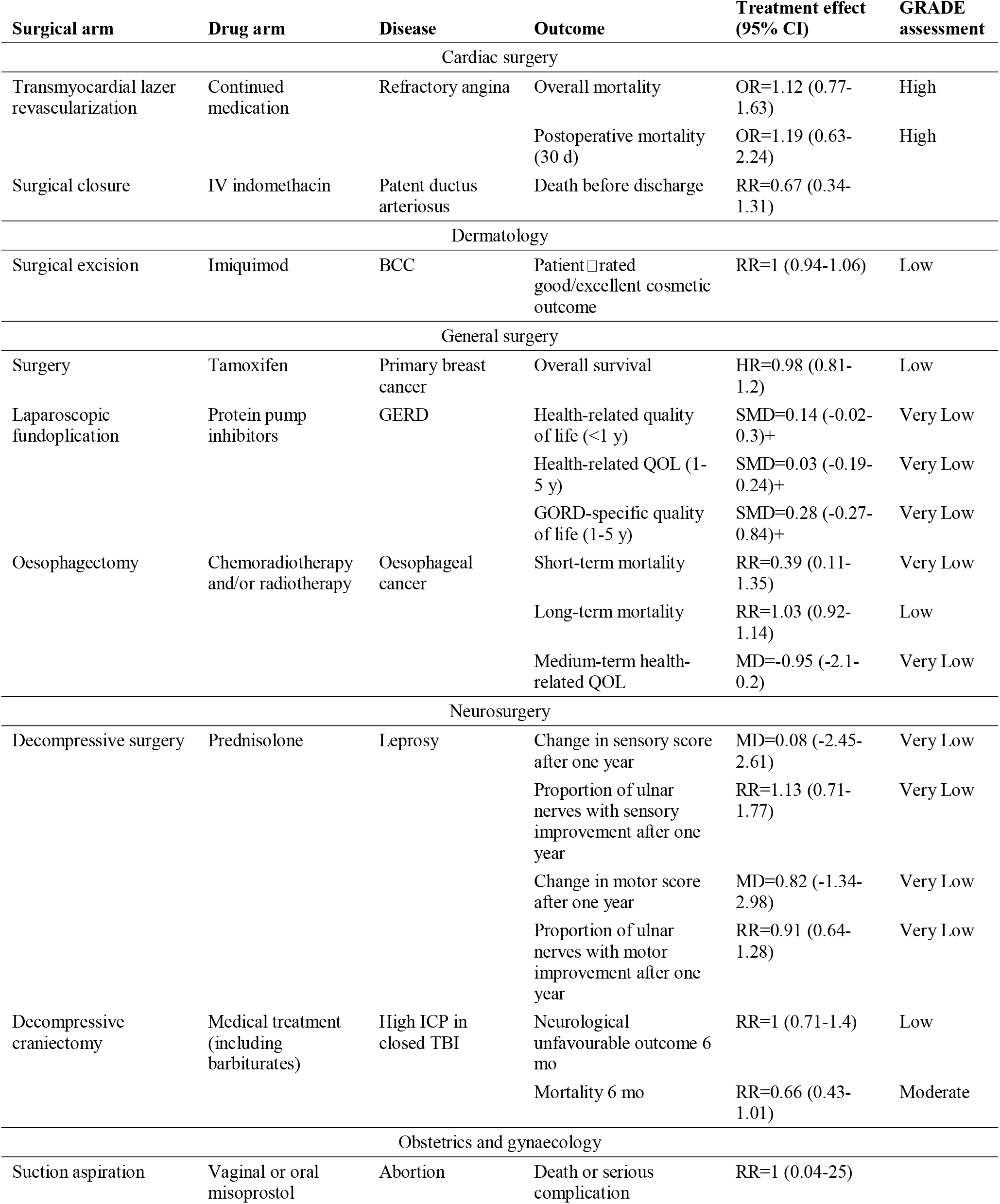
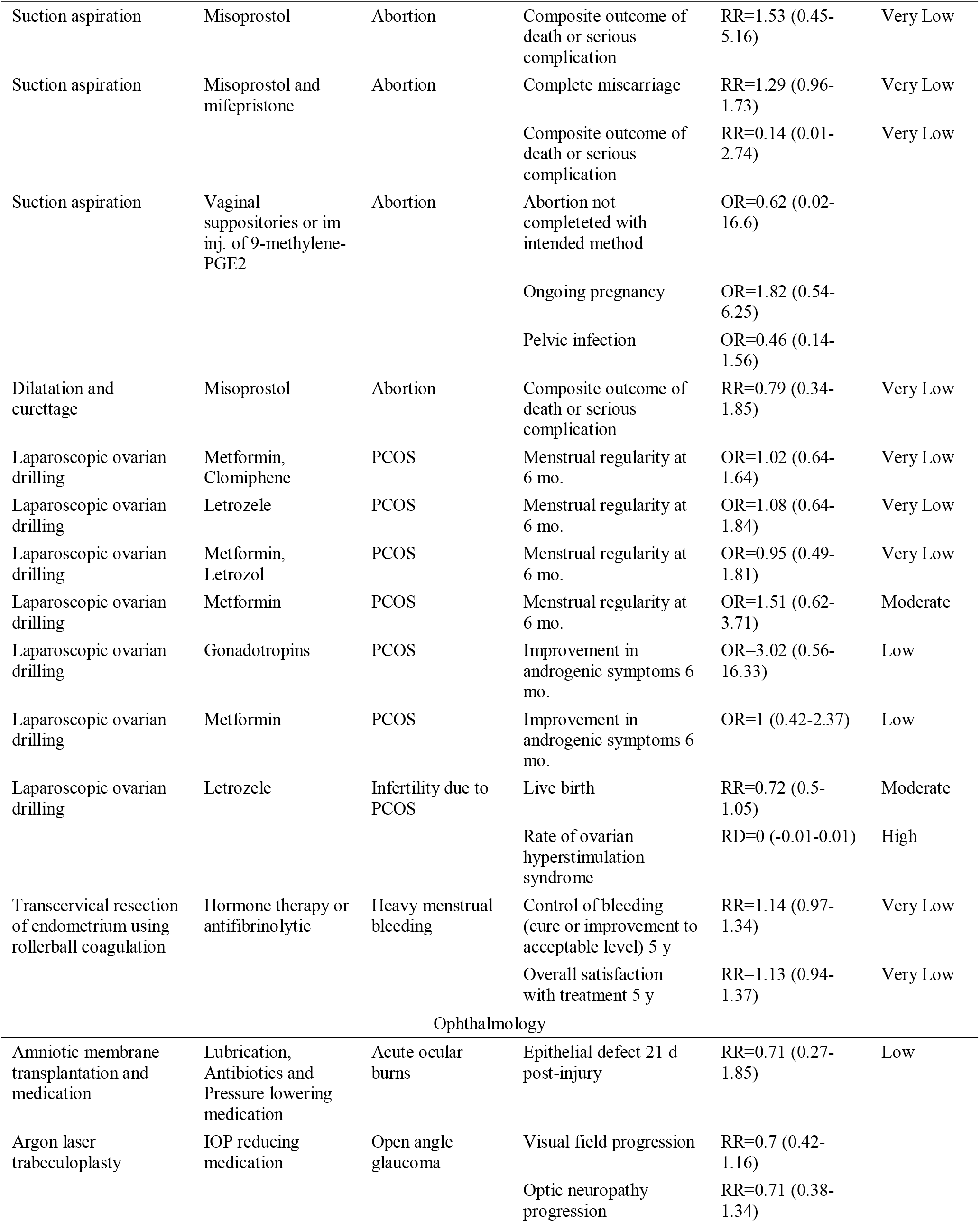
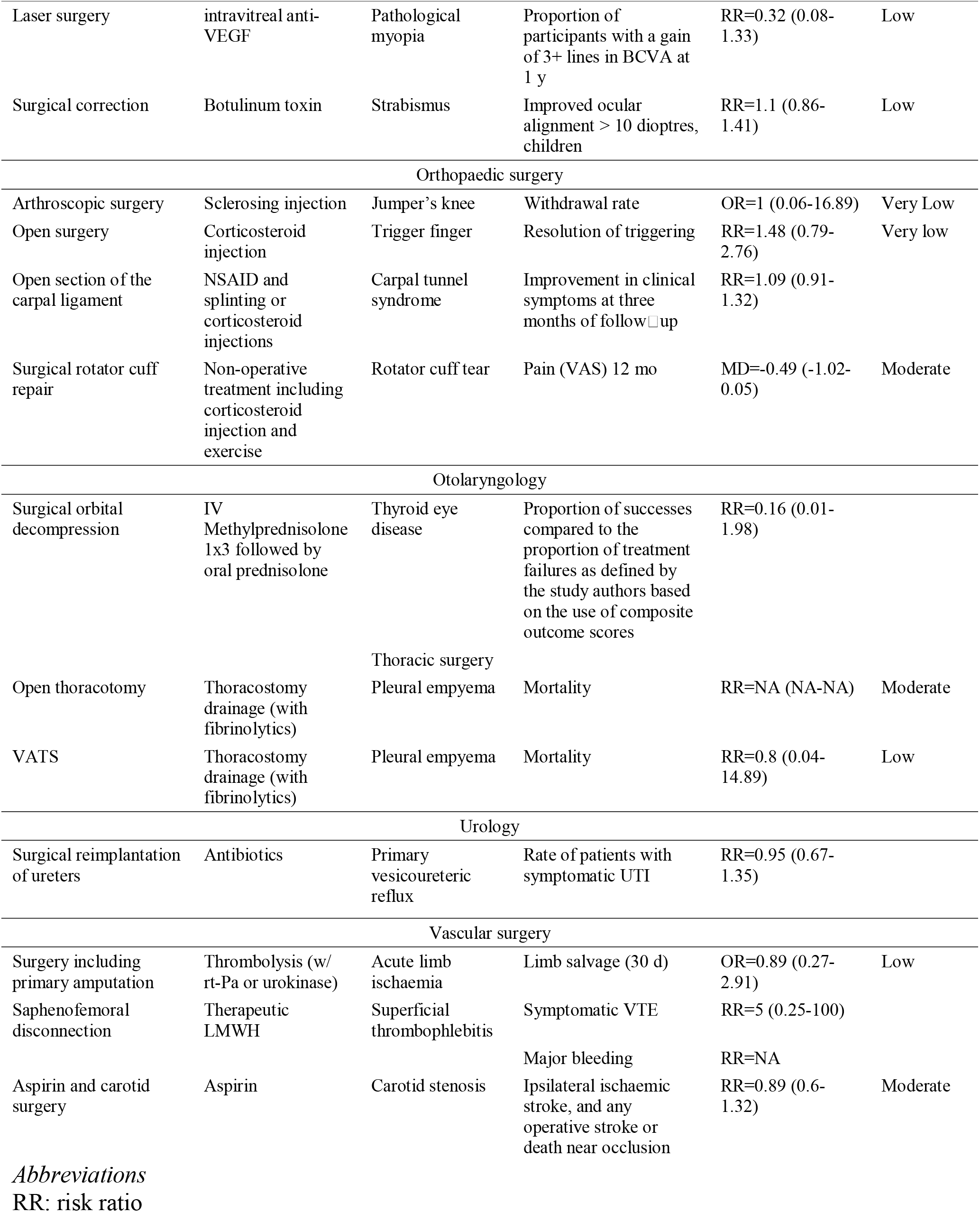
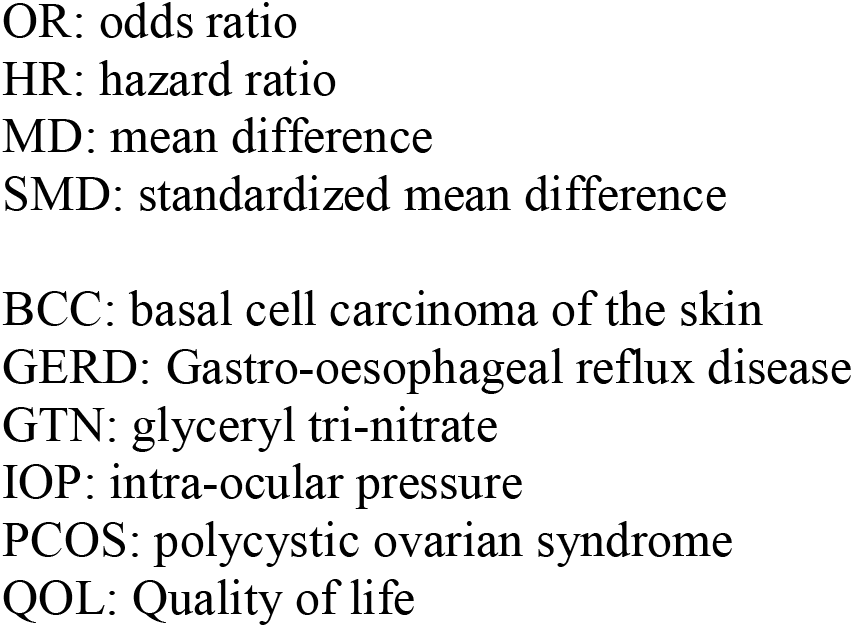
Inconclusive comparisons between surgery and drugs

